## Supplementary information for "Using high-resolution contact networks to evaluate SARS-CoV-2 transmission and control in large-scale multi-day events"

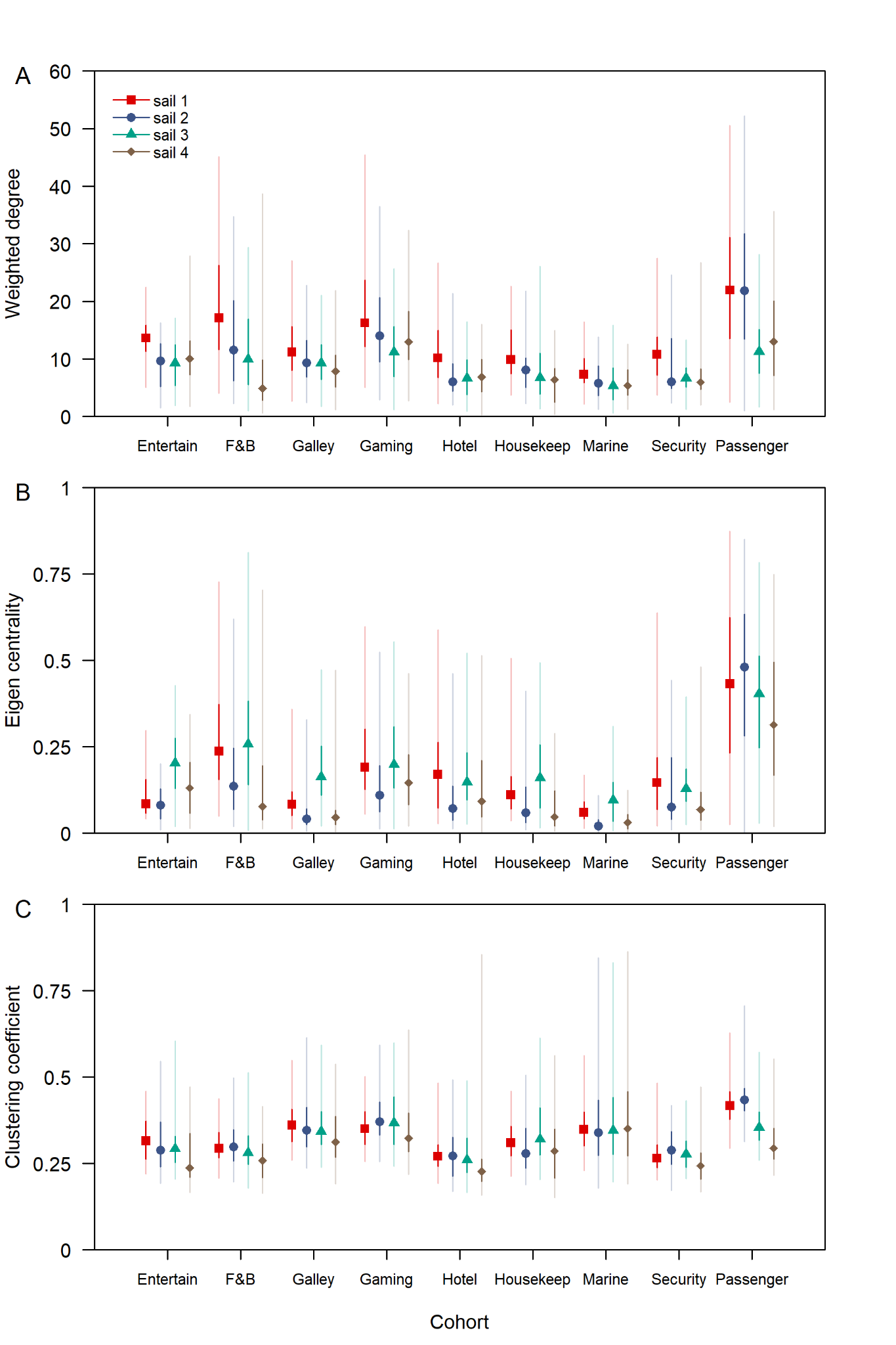

**Fig. S1** Social network analysis over four cruise sailings. (A) Weighted degree, (B) eigenvector centrality, (C) clustering coefficient of crew and passengers of each sailing. Colours represent the cruise departure date and the median (shapes), 50% (dark lines) and 95% intervals (light lines) are shown. Weights were assigned based on exponent transformation of the mean daily cumulative duration of interaction between two individuals (see Materials and methods)

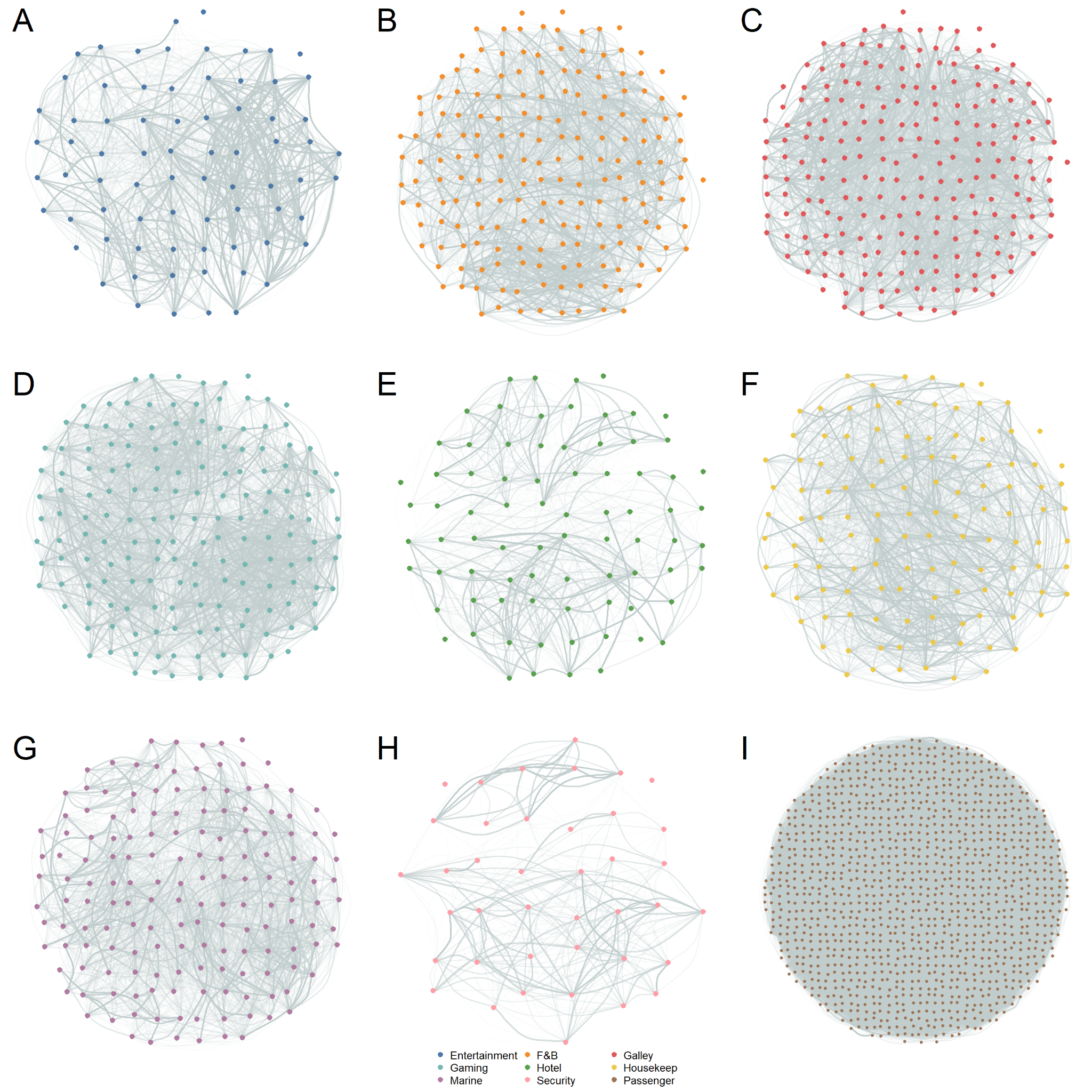

**Fig. S2** Static intra-cohort contacts throughout the entire sailing, with crew from entertainment (A), F&B (B), galley (C), gaming (D), hotel services (E), housekeeping (F), marine (G), security and surveillance (H) departments and passengers (I). In addition, there were 77,107 unique pairs of crew contacts from different cohorts and 70,360 unique pairs of crew and passenger contacts but these links were not represented in this figure. Edge width and colour intensity of the edges correspond to the weights of a contact with the highest colour intensity as shown in the legend. Edge weights are a function of the proportion of days with recorded contact over a three-day sail period and the exponent transformation of the mean daily cumulative contact duration between two individuals.

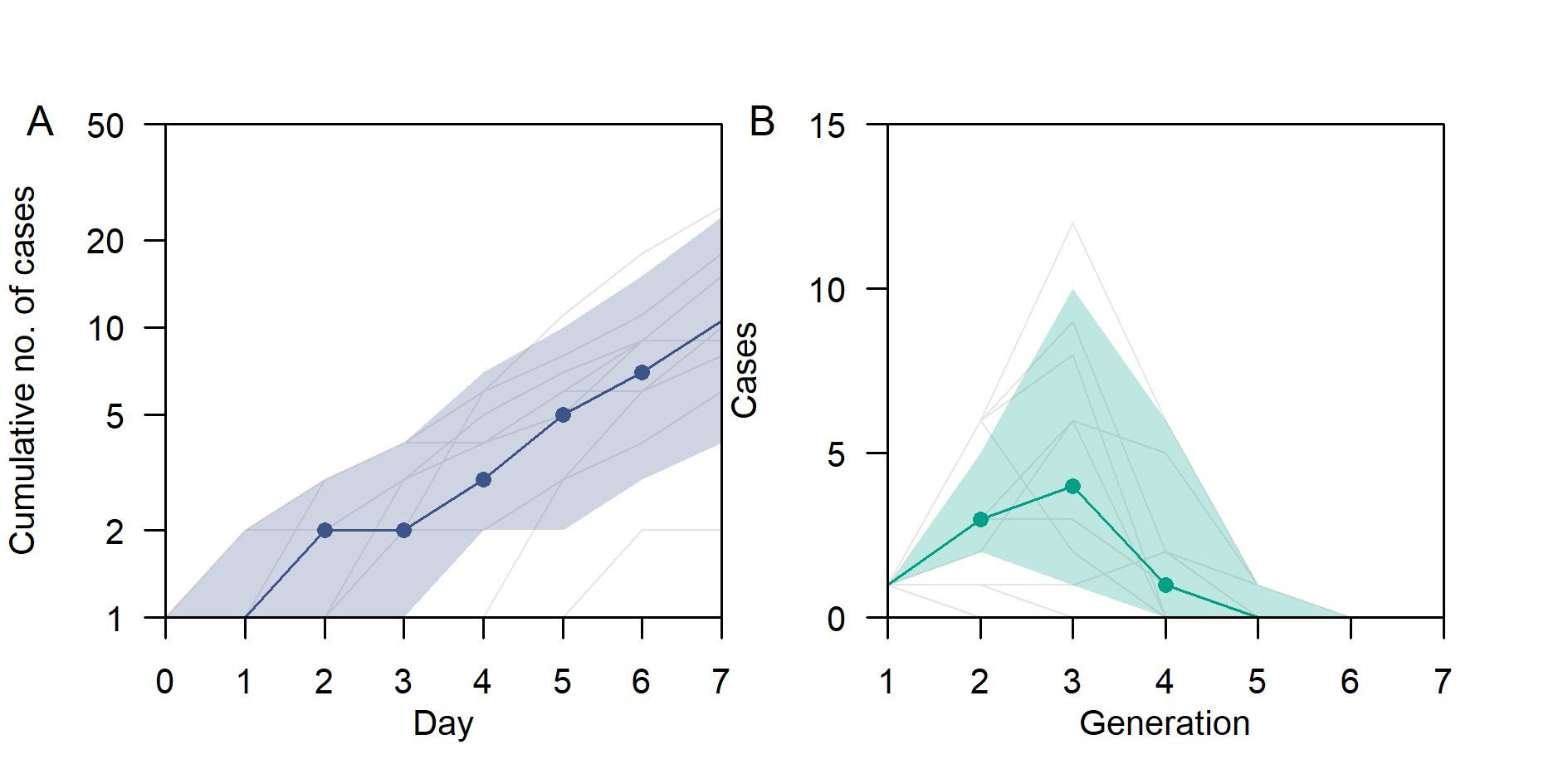

**Fig. S3** (A) Cumulative cases by day of exposure and (B) number of cases in respective generations in the baseline scenario. Median (dots) and 95% intervals (shaded region) and outbreak trajectory for 10 selected simulations (grey lines) are shown.

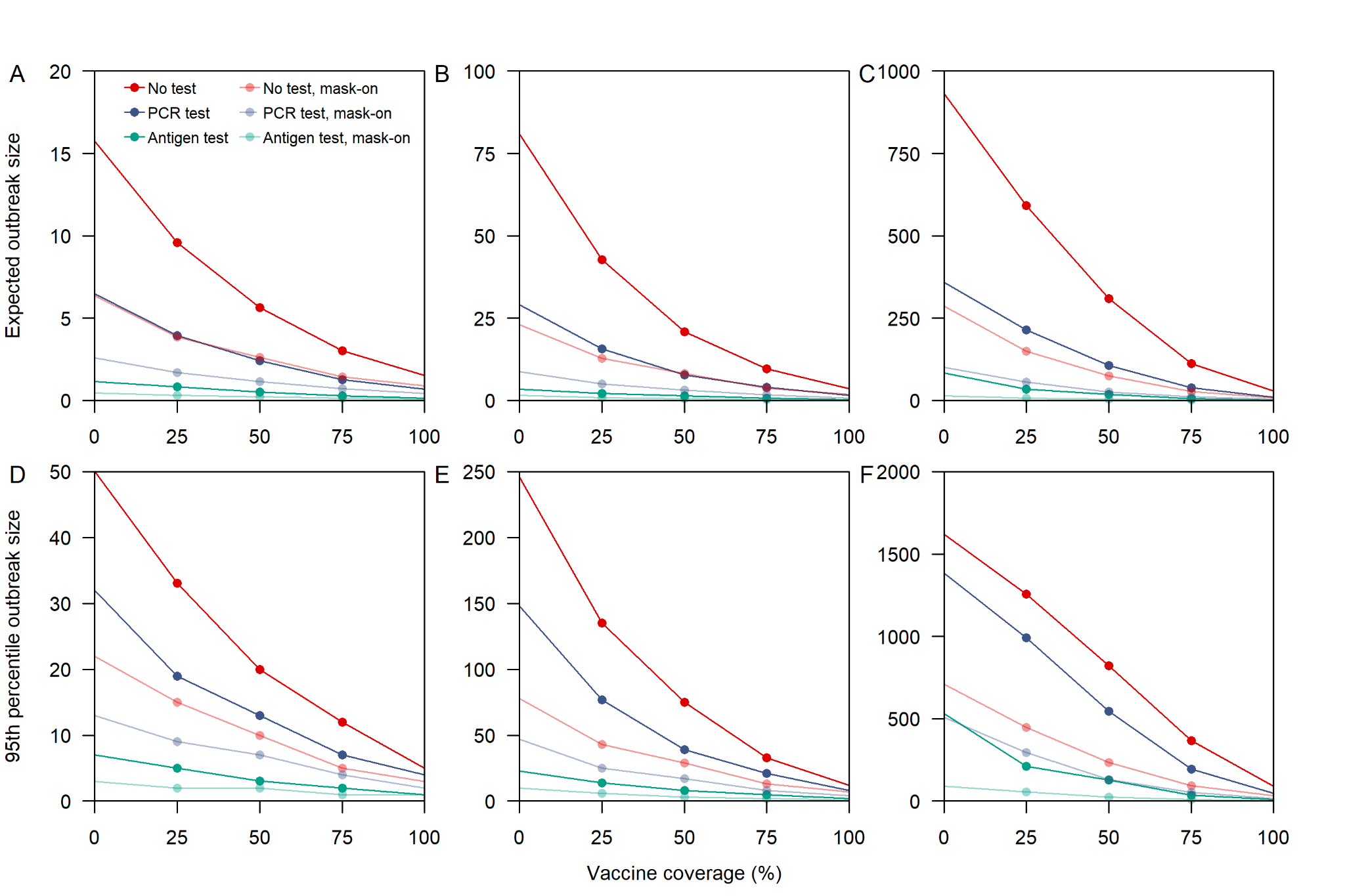

**Fig. S4** Average and 95^th^ percentile in outbreak size for varying interventions, vaccination coverage and assumption on network edge. Vaccines were assumed to confer 50% protection against infection and 50% lowered infectiousness for breakthrough infections in vaccinated individuals. Presymptomatic transmission was modelled to occur in 25% of the infections. (A, D) Edge weights vary based on the proportion of days with recorded interaction over a three-day sail period and duration of contact with weights increasing with days of interaction and contact time but reaches 95% saturation after 3 hours of contact, (B, E) same as (A, D) but reaches 95% saturation after 1 hour of contact, (C, F) edge weights vary based on proportion of days with recorded interaction.

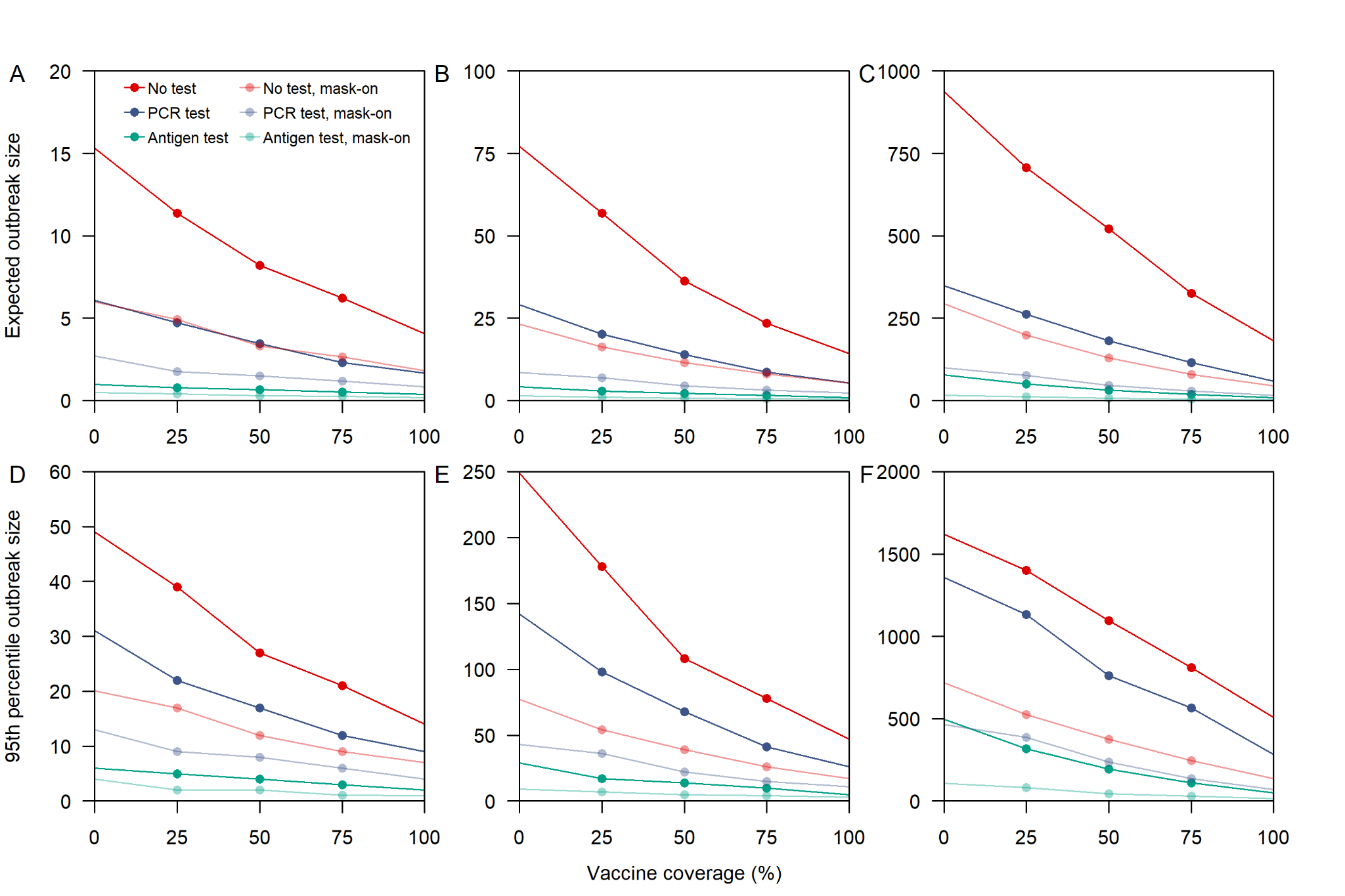

**Fig. S5** Average and 95^th^ percentile in outbreak size for varying interventions, vaccination coverage and assumption on network edge. Vaccine was assumed to confer 50% protection against infection but no lowered infectiousness. Presymptomatic transmission was modelled to occur in 25% of the infections. (A, D) Edge weights vary based on the proportion of days with recorded interaction over a three-day sail period and duration of contact with weights increasing with days of interaction and contact time but reaches 95% saturation after 3 hours of contact, (B, E) same as (A, D) but reaches 95% saturation after 1 hour of contact, (C, F) edge weights vary based on proportion of days with recorded interaction.

Relative to supplementary fig. 4, the expected outbreak size of all simulations increased across all vaccination coverage but the trend of outbreak size across varying coverage and differences between different combinations of interventions remains relatively unchanged

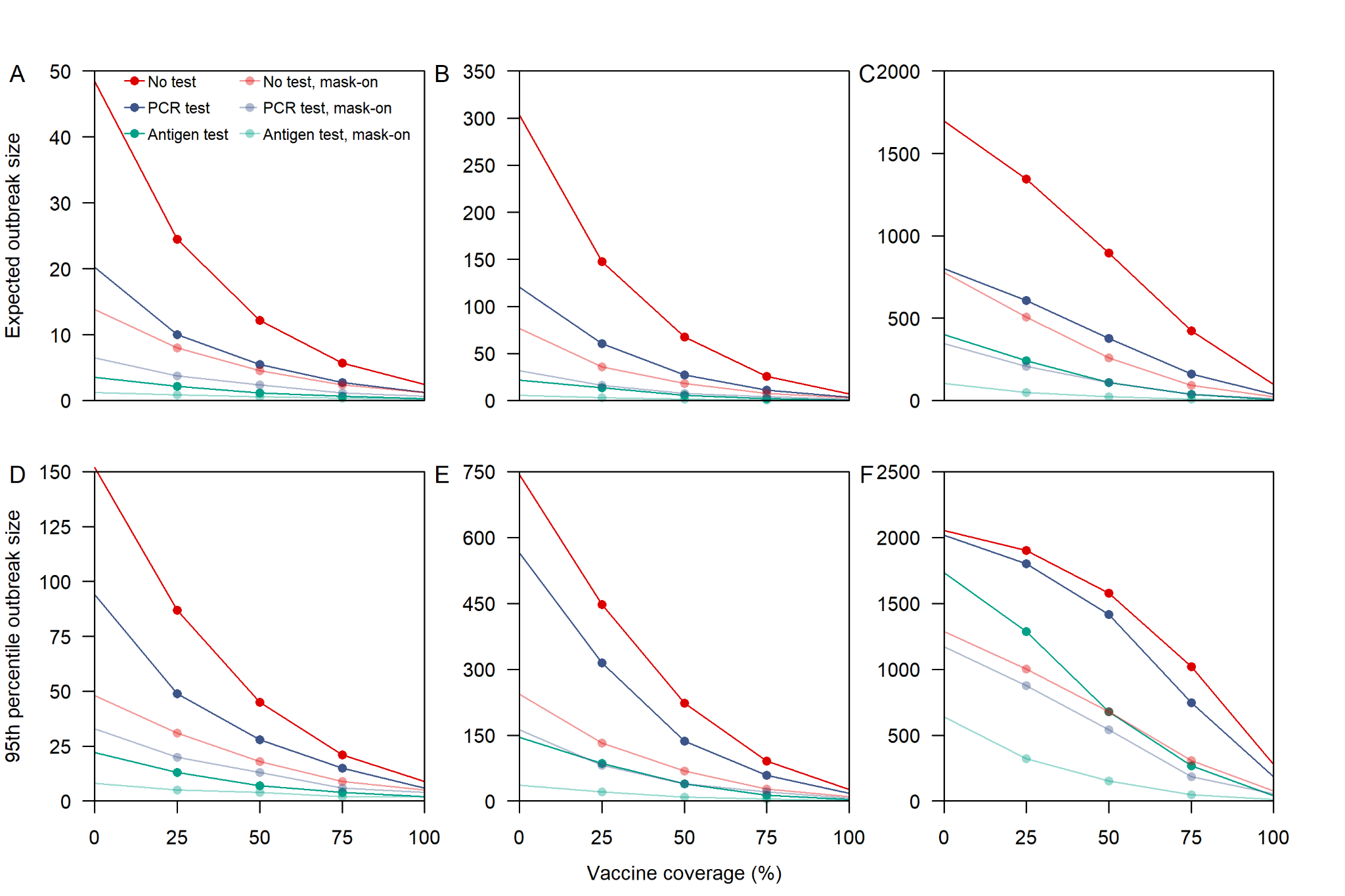

**Fig. S6** Average and 95^th^ percentile in outbreak size for varying interventions, vaccination coverage and assumption on network edge. Vaccine was assumed to confer 50% protection against infection and 50% lowered infectiousness. Presymptomatic transmission was modelled to occur in 50% of the infections. (A, D) Edge weights vary based on the proportion of days with recorded interaction over a three-day sail period and duration of contact with weights increasing with days of interaction and contact time but reaches 95% saturation after 3 hours of contact, (B, E) same as (A, D) but reaches 95% saturation after 1 hour of contact, (C, F) edge weights vary based on proportion of days with recorded interaction.

Relative to supplementary fig. 4, individuals with onset late into the event were able to generate more infections and drove up the expected outbreak sizes. Furthermore, the differences between a mask-off, once off PCR intervention and a mask-on baseline intervention widens with the former having lowered potential in identifying cases prior to the event. At low or no vaccine coverage, the 95th percentile outbreak size under mask-on interventions was lower than that for mask-off interventions with the latter approaching an outbreak size of 90%.

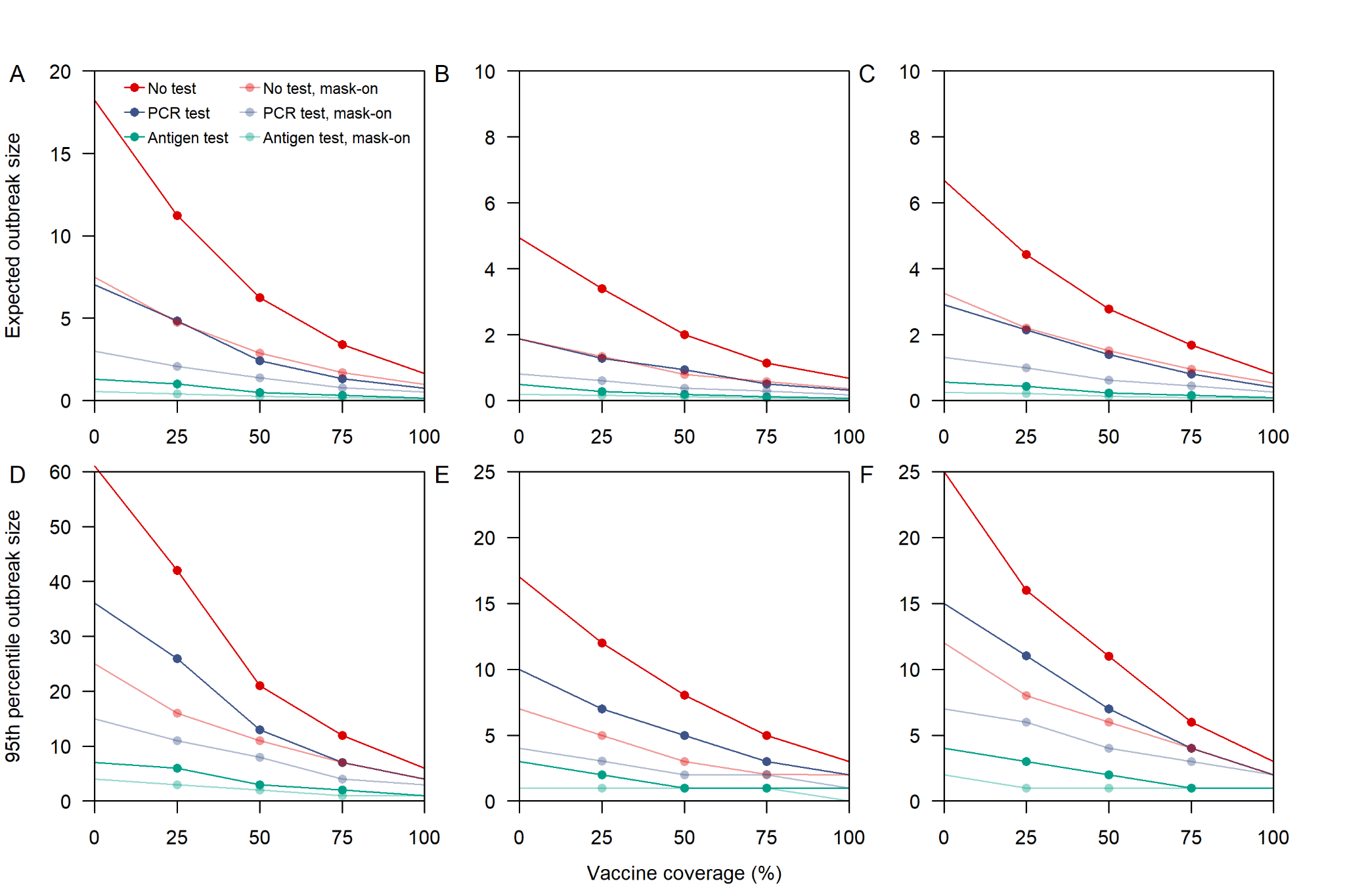

**Fig. S7** Average and 95^th^ percentile in outbreak size for varying interventions, vaccination coverage and for different cruise sailings on second (A,D), third (B,E) and fourth (C,F) sailing. Vaccine was assumed to confer 50% protection against infection and 50% lowered infectiousness. Presymptomatic transmission was modelled to occur in 25% of the infections. Edge weights vary based on the proportion of days with recorded interaction over a three-day sail period and duration of contact with weights increasing with days of interaction and contact time but reaches 95% saturation after 3 hours of contact.

Table S1.

Work functions of respective crew department

| Department | Work functions |
| --- | --- |
| Entertainment | Cruise shows, live entertainment |
| Food & Beverage | From-end consumer facing food and beverages services |
| Galley | Back-end non-consumer facing galley, provision, stewarding |
| Gaming | Casinos, sports, arcade |
| Hotel | Hotel admin, front desk, embarkation training, spa, finance, IT, retail |
| Housekeep | Housekeeping, laundry |
| Marine | Deck, safety, security, medical, engineers, technicians, contractors |
| Security | Surveillance, security |

**Supplementary Text 1:** Singapore CruiseSafe working group

The following authors were part of the Singapore CruiseSafe working group. Each contributed in the collection of data, development of policies for safe resumption of cruise, execution of the operations, contributed to the manuscript, and approved the work for publication: (Singapore Tourism Board) Annie Chang, Jade Kong, Jazzy Wong, Ooi Jo Jin, (Ministry of Health, Singapore) Deepa Selvaraj, Dominique Yong, Jocelyn Lang, (Government Technology Agency) Abilash Sivalingam.

**Supplementary Text 2:** CMMID COVID-19 Working Group members and funding

The following authors were part of the Centre for Mathematical Modelling of Infectious Disease COVID-19 Working Group. Each contributed in processing, cleaning and interpretation of data, interpreted findings, contributed to the manuscript, and approved the work for publication: Simon R Procter, Stefan Flasche, William Waites, Kiesha Prem, Carl A B Pearson, Hamish P Gibbs, Katharine Sherratt, C Julian Villabona-Arenas, Kerry LM Wong, Yang Liu, Paul Mee, Lloyd A C Chapman, Katherine E. Atkins, Matthew Quaife, James D Munday, Sebastian Funk, Rosalind M Eggo, StÈphane HuÈ, Nicholas G. Davies, David Hodgson, Kaja Abbas, Ciara V McCarthy, Joel Hellewell, Sam Abbott, Nikos I Bosse, Oliver Brady, Rosanna C Barnard, Mark Jit, Damien C Tully, Graham Medley, Fiona Yueqian Sun, Christopher I Jarvis, Rachel Lowe, Kathleen O'Reilly, Sophie R Meakin, Akira Endo, Frank G Sandmann, W John Edmunds, Mihaly Koltai, Emilie Finch, Amy Gimma, Alicia Rosello, Billy J Quilty, Yalda Jafari, Gwenan M Knight, Samuel Clifford, Timothy W Russell.

The following funding sources are acknowledged as providing funding for the working group authors. This research was partly funded by the Bill & Melinda Gates Foundation (INV-001754: MQ; INV-003174: KP, MJ, YL; INV-016832: SRP; NTD Modelling Consortium OPP1184344: CABP, GFM; OPP1139859: BJQ; OPP1191821: KO'R). BMGF (INV-016832; OPP1157270: KA). CADDE MR/S0195/1 & FAPESP 18/14389-0 (PM). EDCTP2 (RIA2020EF-2983-CSIGN: HPG). ERC Starting Grant (#757699: MQ). ERC (SG 757688: CJVA, KEA). This project has received funding from the European Union's Horizon 2020 research and innovation programme - project EpiPose (101003688: AG, KLM, KP, MJ, RCB, WJE, YL). FCDO/Wellcome Trust (Epidemic Preparedness Coronavirus research programme 221303/Z/20/Z: CABP). This research was partly funded by the Global Challenges Research Fund (GCRF) project 'RECAP' managed through RCUK and ESRC (ES/P010873/1: CIJ). HDR UK (MR/S003975/1: RME). HPRU (This research was partly funded by the National Institute for Health Research (NIHR) using UK aid from the UK Government to support global health research. The views expressed in this publication are those of the author(s) and not necessarily those of the NIHR or the UK Department of Health and Social Care200908: NIB). MRC (MR/N013638/1: EF; MR/V027956/1: WW). Nakajima Foundation (AE). NIHR (16/136/46: BJQ; 16/137/109: BJQ, FYS, MJ, YL; 1R01AI141534-01A1: DH; NIHR200908: LACC, RME; NIHR200929: CVM, FGS, MJ, NGD; PR-OD-1017-20002: AR, WJE). Royal Society (Dorothy Hodgkin Fellowship: RL). UK DHSC/UK Aid/NIHR (PR-OD-1017-20001: HPG). UK MRC (MC_PC_19065 - Covid 19: Understanding the dynamics and drivers of the COVID-19 epidemic using real-time outbreak analytics: NGD, RME, SC, WJE, YL; MR/P014658/1: GMK). UKRI (MR/V028456/1: YJ). Wellcome Trust (206250/Z/17/Z: TWR; 206471/Z/17/Z: OJB; 208812/Z/17/Z: SC, SFlasche; 210758/Z/18/Z: JDM, JH, KS, SA, SFunk, SRM; 221303/Z/20/Z: MK). No funding (DCT, SH).
